# Effects of environmental factors on severity and mortality of COVID-19

**DOI:** 10.1101/2020.07.11.20147157

**Authors:** Domagoj Kifer, Dario Bugada, Judit Villar-Garcia, Ivan Gudelj, Cristina Menni, Carole Sudre, Frano Vučković, Ivo Ugrina, Luca F Lorini, Margarita Posso, Silvia Bettinelli, Nicola Ughi, Alessandro Maloberti, Oscar Epis, Cristina Giannattasio, Claudio Rossetti, Livije Kalogjera, Jasminka Peršec, Luke Ollivere, Benjamin J Ollivere, Huadong Yan, Ting Cai, Guruprasad P. Aithal, Claire J Steves, Anu Kantele, Mikael Kajova, Olli Vapalahti, Antti Sajantila, Rafal Wojtowicz, Waldemar Wierzba, Zbigniew Krol, Artur Zaczynski, Katarina Zycinska, Marek Postula, Ivica Lukšić, Rok Čivljak, Alemka Markotić, Johannes Brachmann, Andreas Markl, Christian Mahnkopf, Benjamin Murray, Sebastien Ourselin, Ana M. Valdes, Juan P Horcajada, Xavier Castells, Julio Pascual, Massimo Allegri, Dragan Primorac, Tim D. Spector, Clara Barrios, Gordan Lauc

## Abstract

**Background:** Most respiratory viruses show pronounced seasonality, but for SARS-CoV-2 this still needs to be documented.

**Methods:** We examined the disease progression of COVID-19 in 6,914 patients admitted to hospitals in Europe and China. In addition, we evaluated progress of disease symptoms in 37,187 individuals reporting symptoms into the COVID Symptom Study application.

**Findings:** Meta-analysis of the mortality risk in seven European hospitals estimated odds ratios per one day increase in the admission date to be 0.981 (0.973-0.988, p<0.001) and per increase in ambient temperature of one degree Celsius to be 0.854 (0.773-0.944, p=0.007). Statistically significant decreases of comparable magnitude in median hospital stay, probability of transfer to Intensive Care Unit and need for mechanical ventilation were also observed in most, but not all hospitals. The analysis of individually reported symptoms of 37,187 individuals in the UK also showed the decrease in symptom duration and disease severity with time.

**Interpretation:** Severity of COVID-19 in Europe decreased significantly between March and May and the seasonality of COVID-19 is the most likely explanation.

## Background

Over a million of COVID-19 related deaths have been reported until October 1^st^ 2020, but a significant number of people (over 80% in some populations) infected with SARS-CoV-2 manage to contain infection in their upper respiratory tract and despite being PCR positive for the viral RNA do not develop any visible symptoms (1). So far, very little attention has been given to the effects of environmental conditions on the individual course of the diseases.

The first study of the environmental effects on the COVID-19 infection rate in 30 Chinese provinces found significant negative associations with temperature and relative humidity in Hubei province with the decrease of cases by 36%-57% for every 1 °C and 11%-22% for every 1% increase in relative humidity, these associations were inconsistent in other provinces (2). Negative effects on COVID-19 transmission with warmer temperatures were also observed in Turkey (3), Mexico (4), Brazil (5) and United States (6), while similar association with humidity was reported in Australia, but with temperature having no effect on the virus transmission (7). The study from Brazil observed flattening of the temperature effect on the virus transmission at 25.8°C thus suggesting that warmer weather will not cause the transmission decline which is in accordance with the studies from Iran and Spain where they observed no changes in transmission rates under different temperatures and humidity (8,9). These studies are inconsistent and do not give clear evidence as to whether there is an association between the temperature, humidity and virus transmission, the global view seems to give a clearer conclusion; all the three studies which conducted analysis at the global level found an association between higher humidity, warmer temperatures and lower transmission rate (10). However, climate-dependent epidemic modelling suggested that the absence of population immunity is a much stronger factor in viral transmission and that summer weather will not substantially limit the spread of COVID-19 pandemics (11). This is consistent with high numbers of infected individuals in tropical countries and the increase of cases in the south of United States in the second half of June 2020.

Recent studies report increasing numbers of SARS-Cov2 positive asymptomatic individuals (1), but it is not clear whether the apparent increase in people with mild or no symptoms is due to the change in the extent of testing, or some other characteristic of the SARS-CoV-2 virus. Aiming to evaluate the association of humidity and ambient temperature with the severity of the COVID-19 disease, we analysed individual-patient data for 6,914 patients with COVID-19 admitted to hospitals in Bergamo, Italy: Barcelona, Spain; Coburg, Germany; Helsinki, Finland; Milan, Italy; Nottingham, United Kingdom; Warsaw, Poland; Zagreb, Croatia and Zhejiang province, China since the beginning of the pandemics and compared it to environmental temperature and calculated indoor humidity. Furthermore, we analysed information about COVID-19 severity from the COVID Symptom Study application that is collecting information of 37,187 individuals in the UK.

## Methods

### Studied cohorts

We collected information about hospital admission, discharge dates, admission to intensive care unit (ICU), need for mechanical ventilation and type of discharge (alive or dead) for 5229 successive patients hospitalized for COVID-19 in six European Hospitals and 13 hospitals in Zhejiang province, China since the beginning of the pandemics (Table 1). We included patients with confirmed diagnosis of COVID-19 at the time of admission. We confirmed that patients had a positive result on polymerase chain reaction testing of a nasopharyngeal sample and/or a clinically/radiologically diagnosis of COVID-19. Patients were not followed after discharge, but COVID-19 related early readmissions were considered as part of the COVID-19 course. The study protocol conformed to the ethical guidelines of the 1975 Declaration of Helsinki. In Zhejiang hospitals, ASST Papa Giovanni XXIII° Hospital in Bergamo, Hospital del Mar in Barcelona and Helsinki University Hospital local ethics committees approved this retrospective study of COVID-19 patient data. For REGIOMED Hospital in Coburg, Ethics committee of the Bavarian state physician’s association approved the study. In Nottingham University Hospital’s trust, ASST GOM Niguarda, Warsaw and Zagreb this information was released as public statistical information.

**Table 1.** Basic information about included patients

| Cohort name | Hospital name | Total number of patients | Included in the study | Sex (f/m) | Age (median, range) |
| --- | --- | --- | --- | --- | --- |
| Barcelona | Hospital del Mar | 1999 | 1786 | 969 / 817 | 57 (17 – 101) |
| Bergamo | ASST Papa Giovanni XXIII<sup>o</sup> Hospital | 2249 | 995 | 265 / 730 | 70 (6 – 95) |
| Coburg | REGIOMED | 89 | 89 | 48 / 41 | 75 (18 – 98) |
| Helsinki |  | 100 | 100 | 44 / 56 | 54.5 (16 – 84) |
| Milan | Asst GOM Niguarda | 713 | 685 | 242 / 443 | 63 (0 – 96) |
| Nottingham | Nottingham University Hospitals | 795 | 795 | 356 / 439 | 75 (0 – 102) |
| Warsaw | Central Clinical Hospital of Ministry of the Interior and Administration | 122 | 122 | 45 / 77 | 69 (19 – 96) |
| Zagreb | Clinical Hospital Dubrava & University Hospital for Infectious Diseases | 237 | 237 | 93 / 144 | 63 (22 – 99) |
| Zhejiang |  | 610 | 608 | 297 / 311 | 49 (18 – 93) |

### COVID Symptom Study application

The COVID Symptom Study app(12) developed by Zoe with scientific input from researchers and clinicians at King’s College London and Massachusetts General Hospital, (https://covid.joinzoe.com/) was launched in the UK on Tuesday the 24th March 2020, and in 3 months reached more than 3.9 million subscribers. It enables capture of self-reported information related to COVID-19 infections, as reported previously (12). Importantly, participants enrolled in ongoing epidemiologic studies, clinical cohorts, or clinical trials, can provide informed consent to link data collected through the app in a HIPPA and GDPR-compliant manner with extant study data they have previously provided or may provide in the future. The Ethics for the app has been approved by King’s College London ethics Committee (REMAS ID 18210, review reference LRS-19/20-18210) and all users provided consent for non-commercial use. For this work we included participants from the United Kingdom who started reporting with a healthy status and subsequently developed symptoms leading to suspect COVID-19 following the disease score presented in Menni et al. (12) In order to get an estimate of disease duration, the time for disease end corresponded to either the last day of report before stopping using the app, or the first healthy day when followed by 6 consecutive days of healthy reporting. To avoid censoring, only participants with a disease duration of less than 30 days and with a disease onset occurring before the 17^th^ May were included in the analysis (37,187 individuals). Severity score was calculated as a weighted average of symptoms at disease peak using as weight the normalised ratio in symptom frequency at disease peak between people reporting hospital visit after disease onset and those that did not.

### Data related to seasonal changes

Ambient temperature data was obtained from the Climate Data Online (National Centers for Environmental Information (NCEI) database): https://www.ncdc.noaa.gov/cdo-web/

### Statistical Methods

The data collated from 7 cohorts are summarised in Table 1. Patients without information about outcome were excluded from the analysis. Logistic regression was used to estimate the effect of admission date and local ambiental temperature on mortality change. The following patient characteristics, and hospitalization episode co-variates were explored: Died/discharged outcome was used as dependent variable and admission as independent variable along with age (in years) and gender (female/male). We them used the same approach for estimating the effect of ambient temperature on need for admission to ICU, and for mechanical ventilation therapy. A linear model was then used to estimate the effect of ambient temperature on the hospital stay length (in days) as dependent variable, and admission date as independent variable along with age and gender. Prior the analysis data transformation was undertaken with hospital length of stay increased by 1 (due to zeros) and log10 transformed (Zero days in hospital stay correspond to hospitalization with a length lower than 24 hours). Linear regression using median duration as dependent variable and 2-week period as independent variable was fitted to assess change over time.

For each dependent variable, raw data were presented with bar plots (death, ICU and mechanical ventilation) or box-and-whiskers plots (hospital length of stay) for patients in two-week groups. Fill of bars and boxes reflects the number of patients admitted to hospital in particular two-week group. With groups of less than 5 patients individual data points were plotted.

Coefficients estimated in logistic regressions and linear regression were combined using an inverse variance-weighted meta analyses methods where given the heterogenity of cohorts random effects models were used (R package “metaphor”).

Results of the meta-analysis were presented as forest plots, created using R package “ggplot2”. All statistical analyses were performed in R programming software (version 3.6.3), with exception of logistic and linear regressions on Milano cohort data which are performed in Stata Statistical Software (version 12) and the COVID Symptom Study cohort for which linear regression were performed using python statsmodels package (version 0.11.1).

## Results

Aiming to evaluate seasonal nature of COVID-19, we evaluated disease course in 6,914 individuals from nine cohorts admitted to hospitals in Europe and China (Table 1). To avoid sampling bias, all hospitalizations that resulted in either death, or medical discharge were included in the analysis. Actual numbers of patients who died and patients who recovered (grouped in two-week intervals) since the beginning of the epidemics, until the final follow up date for reliable data capture reporting final outcome was available are presented in Figure 1A for each of the hospitals. Meta-analysis of the effect of admission date on the mortality is presented in Figure 1B. The most significant change was observed in Barcelona, where mortality odds decreased by 4.1% per day (p<0.001). Weighted average decrease in mortality odds across all studied hospitals was 1.9% per day (p<0.001). Our model included age as a co-variate, so this change is unlikely to be accounted for by change in age of patients. To further confirm that age was not underlying the observed changes we analysed age of patients admitted to hospitals in different periods and demonstrated that change in the age of patients was not a factor that could explain the observed decrease in mortality (Supplementary Figure 1).

**Figure 1.**
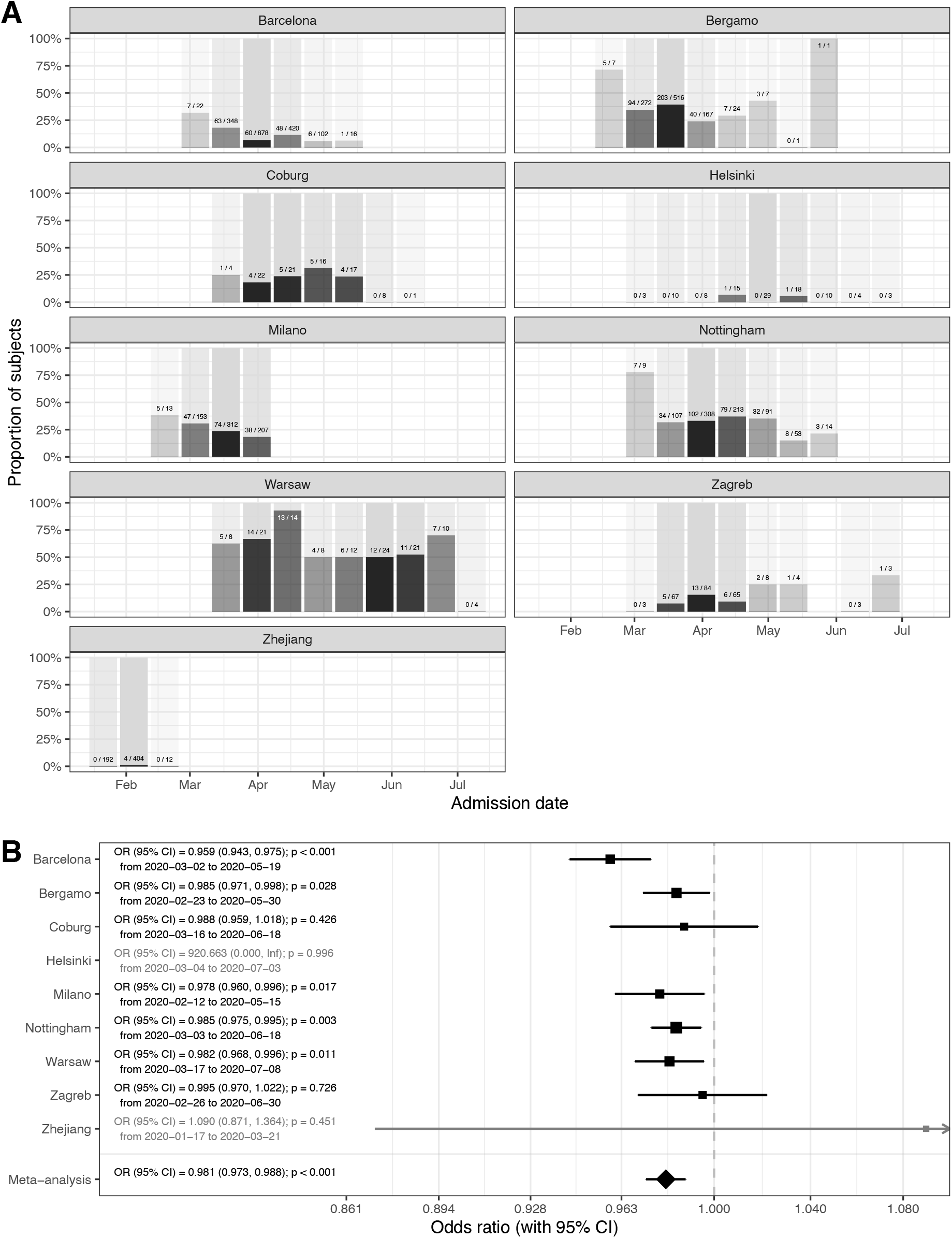
Mortality in people admitted in hospitals with COVID-19. A – hospitalization outcome (death/discharge) depending on the admission date (grouped in two-week intervals) since the beginning of the pandemics; B - Meta-analysis of the effects of admission date on the mortality (presented as odds ratios per one day increase in admission date). In Helsinki there were only 2 deaths and in Zhejiang hospitals 4 deaths, so they were not included in the meta-analysis. OR – odds ratio, CI – confidence interval.

Since there is no standard measure or classification of COVID-19 severity used across all hospitals, to further evaluate disease severity, we analysed secondary outcomes. We compared the duration of hospitalization, need for intensive care unit (ICU) and separately mechanical ventilation. Strong and statistically significant decrease in the duration of hospitalization was observed in Barcelona, Coburg, Milano, Nottingham and Zagreb. In Helsinki, Warsaw and Zhejiang the change was in the same direction but was not statistically significant. The only outlier was Bergamo, where the change was in the opposite direction, but the change was not statistically significant (Supplementary Figure 2). In meta-analysis the decrease in lengths of hospitalization was statistically significant (10^b=0.995; CI=0.991,-0.998); p=0.007). The odds to need of intensive care decreased in all hospitals in Europe and was individually statistically significant in all hospitals beside Bergamo, Helsinki and Zagreb (Supplementary Figure 3). Meta-analysis of European hospitals estimated that the odds to need the intensive care decreased by 2.2% per day of change in the admission date (OR=0.978; CI=0.962-0.993; p=0.008) and the odds to need mechanical ventilation decreased 2.1% (OR=0.979; CI=0.964-0.994; p=0.008) per day of change in the admission date (Supplementary Figure 4).

While all hospitals in Europe were basically displaying the same trend of decreasing COVID-19 severity with time, in Zhejiang hospitals there was either no change, or the changes trended non-significantly in opposite direction to European centres. The most notable difference between COVID-19 pandemics in Europe and in China was that while in China the epidemic was entirely during winter, in Europe it covered both winter and spring periods. To evaluate whether weather was an important factor, we correlated the observed changes with local ambient temperature. Minimal and maximal local temperatures for all hospitals a presented in Supplementary Figure 5. To evaluate whether the change in temperature may have been responsible for the observed changes in disease severity, we modelled mortality with ambient temperature instead of admission date. The results presented in Figure 2 suggest strong effect of ambient temperature on the mortality risk (OR=0.854 per one-degree Celsius; CI=0.773-0.944; p=0.007).

**Figure 2.**
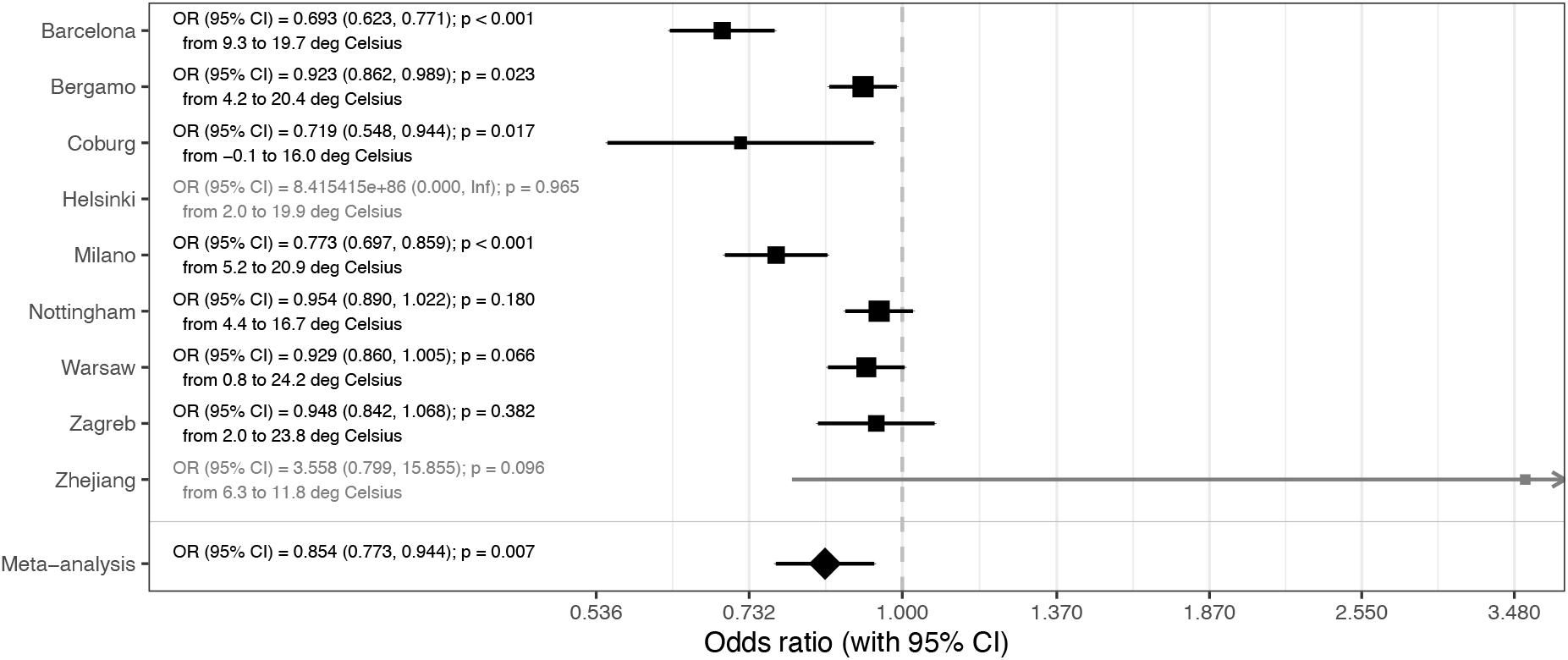
Meta-analysis of the effects of temperature on mortality (presented as odds ratios per one-degree Celsius increase in average daily temperature during hospitalization). In Helsinki there were only 2 deaths and in Zhejiang hospitals 4 deaths, so they were not included in the meta-analysis. OR – odds ratio, CI – confidence interval.

To further verify the change of COVID-19 with time we analysed individual symptom data for 37,187 participants of the Covid Symptom study app. Although there is also a sampling bias in that study, it is a different from bias in hospitalization, so it was reassuring to observe a gradual decrease in duration of symptoms and COVID-19 severity in April and May (Figure 3). An assessment of the slope of duration as function of time (2 ISO week) showed a significant decrease in duration (B=-0.7 p=0.006). Regarding severity, while not overall significant (−0.0014 p=0.836), the trend towards a decrease was stronger when considering the latest period (point 1 to the end slope=-0.0112 p=0.116)

**Figure 3.**
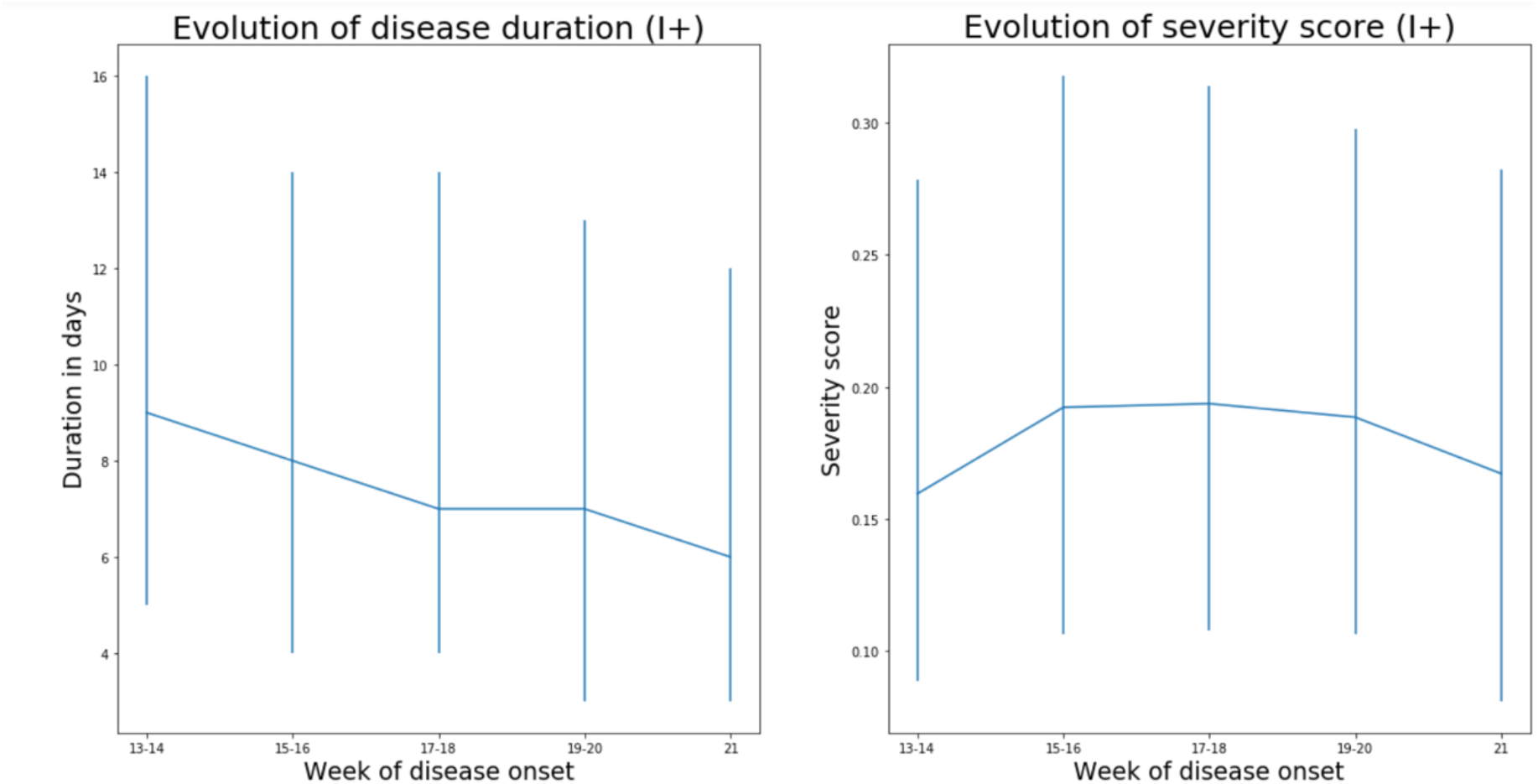
Data from 37,187 individuals suspected COVID positives (I+) recording symptoms in the COVID Symptom Study application in the United Kingdom suggest that both severity (measured as weighted sum of symptoms accounting for difference at disease peak between those reporting hospital visit and those who don’t) and duration of disease symptoms slightly decrease in the United Kingdom. (Median values and interquartile ranges are shown). Imputed status defined as per the application of the predictive model described by Menni et al. (12) was chosen over definite PCR diagnostic in order to avoid confounding factor of test access policy changes.

## Discussion

By analysing hospital records of 6,914 patients admitted to eight European hospitals we observed strong and statistically significant decrease in COVID-19 mortality and severity with time. Possible change in the average age of patients in different stages of the pandemic is the first obvious explanation for the decreased severity, since age is the strongest predictor of COVID-19 severity (with up to 100-fold difference in mortality risk (13)). However, age was included in our model as a co-variate and furthermore the average age of patients did not change with time (Supplementary Figure 2), so we excluded this hypothesis. An alternative explanation could be that there was change in policies for admission and/or release of COVID-19 patients during the evaluated period – possibly due to ‘overwhelming’ of medical facilities. This might have been particularly relevant in the situation of limited hospital capacity, when hospitalization may have been preceded with a triage process to identify patients who might benefit from hospitalization, admission to ICU or mechanical ventilation. However, the only hospital in our cohort that reached full capacity was Bergamo, while all others operated well below the maximal capacity for either hospitalization, or ICU, which suggests that changes in hospital admission policy were not a major driver behind the observed change in COVID-19 mortality and severity. This conclusion is further supported by concurrent decrease in duration and severity of symptoms of non-hospitalized individuals reporting symptoms in the COVID-Symptom Study Application (Figure 3). Change in COVID-19 management, also, could have resulted in decreased severity. However, all these changes were hospital-specific and in the analysed period the most effective improvement in therapy was the introduction of dexamethasone, which was reported to reduce mortality from 24.6% to 21.6% (14). As we are learning more about COVID-19, patients are receiving better and better treatment, but the progress so far was not too large, which is particularly evident from the increased mortality in the second wave in Australia (case fatality rate, CFR was 0.5% in the first wave (15) and 3.1% in the second wave (16)). Therefore, it is hard to imagine that minor modifications in patient management could have significantly contributed to the observed decrease in disease mortality and severity in Europe.

After excluding these three causes for a Europe-wide decrease in disease severity and mortality in the period from March to June, the change in season surfaced as the most probable explanation since in all studied locations ambient temperature increased considerably in that period (Supplementary Figure 5). Exchanging hospital admission date with local temperature (Figure 2) showed that temperature strongly correlated with decrease in COVID-19 mortality. Since reverse causation is not possible, it is reasonable to conclude that COVID-19 as a disease has a strong seasonal nature. Despite the fact that most human coronaviruses are highly seasonal (17), the seasonal nature of COVID-19 is frequently challenged with the fact that numerous cases have been reported in tropical countries and that virus evidently can also be efficiently transmitted in hot and humid climates. However, in all these countries the disease mortality and severity are very low (e.g. Singapore reported 26 deaths and over 44,000 confirmed infections), which actually suggests that there may be seasonal or climate related differences in severity of COVID-19. It is possible that the same is the case for other respiratory viruses that show strong seasonality, but asymptomatic people are generally not tested for the presence of viral RNA in the nose, thus viral transmission, outside of their season, was not observed. The notable exception which confirms this hypothesis was the 2009 swine flu pandemics in England when numerous PCR tests were also performed in the summer. These tests revealed infection in over 250,000 people in the summer wave, but with much lower mortality than in the wither wave (18). The large increase in the number of people with positive SARS-CoV-2 PCR tests in Europe in late summer and early autumn 2020 is not accompanied with the corresponding increase in deaths. The increase of number of cases and change in the age distribution of patients (19) have been suggested as possible explanation for the missing deaths. However, in Australia that has reverse seasons the situation was opposite, and the mortality was much higher in the second (winter) wave of pandemics. Despite increased testing and global increase in the knowledge how to treat patients CFR in the second (winter) wave was six times higher than in the first (summer) wave (“winter” CFR was 3.1% (16) compared to “summer” CFR that was only 0.5% (15)).

It is very difficult to prove causality in an observational study, in particular when many correlated factors are changed in the same time, but the observed decrease in COVID-19 severity with the end of winter fits very well with the known effects of outside temperature on indoor humidity and consequential restoration of mucosal barrier function, which is often impaired by dry air during the heating season (20). Most respiratory viruses peak in winter and fluctuation of temperature and humidity have been proposed as the most potent drivers of seasonality, especially in the context of the epidemics in the winter season (17). However, the peak of infection and the severity of the disease are not always full aligned. For example although infection rates of rhinoviruses peak in spring and fall, the disease severity increases in winter (21). Seasonal appearance of respiratory viruses is often attributed to seasonal indoor crowding and effects of temperature and humidity on stability of viral particles (22), with effect of low air humidity on the mucosal barrier often neglected.

While often considered to be a physical barrier, mucus is actually an active biological barrier that crosslinks viruses and bacteria to mucins, a group of highly glycosylated proteins that are secreted to our mucosal barriers where they self-assemble into long polymers(23). Mucin glycans mimic cell surface glycosylation and by acting as a decoy for viral lectins trap viral particles, which are then transported out of airways by mucociliary clearance(24). Furthermore, since all envelope viruses are highly glycosylated, a number of lectins like trefoil factors (TFF) are secreted to mucous where they crosslink viruses by binding to glycans on both viruses and mucins (25). However, this barrier is functional only if it is well hydrated to both maintain its structural integrity and enable constant flow of mucus that remove viruses and other pathogens from our airways (24). If exposed to dry air, these barriers dry out and cannot perform their protective functions (26).

Animal experiments demonstrated the importance of humidity for both transfection of respiratory viruses and disease severity (27–29), while population-level studies in the United States indicated the importance of humidity for influenza transmission (30). One of these studies demonstrated that increasing relative humidity from 20% to 50% can significantly decrease mortality from influenza infections (29). In another study humidification of air in obstructive sleep apnea patients reduced nasal symptoms by 60% (31), which all suggest that protective effects of humidity on mucosal barrier may be a dominant molecular mechanism behind seasonality of respiratory viruses.

A large part of human inter-individual differences are glycan-based and glycan diversity represent one of the main defences of all higher organisms against pathogens (32). Glycans (which are covalently attached to most proteins) are chemical structures that are being inherited as complex traits, which enables diversity and significant inter-individual differences (33). SARS-CoV-2 spike glycoprotein is heavily glycosylated (34) and it was reported to bind to glycosaminoglycans (35) and sialylated glycans (36). ABO blood antigens are also glycans and are probably the best known example of glycan diversity; interestingly, people with blood type A and, thus, having one N-acetylgalactosamine more than type O are more susceptible to COVID-19. (37). All this suggest that, like most other viruses, SARS-CoV-2 is also dependent on glycans for transmission, which further support the importance of mucins and functional mucosal barrier in COVID-19.

Mucociliary dysfunction and respiratory barrier impairment promotes both initial infection and expansion of viruses within the airways of an infected individual(29). Dry air inhalation significantly decreases nasal mucociliary transition time (NMTT) in heathy individuals(38), affecting the duration of viral exposure on nasal mucosa. Nasal epithelial cells are the main portal for the initial infection and transmission of SARS-CoV-2(39). Patients who developed clinically relevant infection after experimental transnasal viral challenge (Rhinovirus or Infl. B) had reduced epithelial barrier function (increased transepithelial resistance, reduced number of ciliated cells and increased NMTT compared to those who were not infected or had a mild form (40). However, experimental viral infection in vitro resulted only in decreased number of ciliated cells, without affecting tight junction proteins expression (41). This controversy between in vivo and in vitro experiments suggests the importance of immune response in the control of epithelial barrier function (42). Recent studies on the interaction between climate changes and respiratory barrier dysfunction, indicated not only higher incidence of viral infection but also higher vulnerability of nasal mucosa through increased incidence of nosebleed in the emergency departments in the conditions of low temperature and low humidity (43). Recently published study that exposed volunteers to respiratory syncytial virus (RSV), one of the pathogens responsible for the common cold, demonstrated that pre-existing inflammation in the respiratory mucosa was a risk factor for infection (44), which further supports the importance of mucociliary dysfunction and respiratory barrier impairment for infection with respiratory viruses.

### Limitations

Potential sampling bias is the main limitation of this study. By focusing on individual progression of the disease in already hospitalized patients we excluded effects of the unknown number of true infections on national mortality rates, and we still cannot exclude the possibility that some other unidentified external factors (including confinement and social distancing, improvement and compliance of prevention and environmental hygiene protocols and even decreased air-pollution could have progressively affected the severity of patients arriving to the hospital) were affecting composition of hospitalized patient cohorts and contributing to the decreased COVID-19 severity and mortality. Therefore, it is important that tracking of individual symptoms in 37,187 UK patients are showing the same trend, since these are individuals voluntary reporting symptoms and potential sampling bias there is independent from bias in hospitalization. The choice to include imputed positives was mostly motivated by the restriction in testing access that were observed over the first wave before being relaxed in May and June. Accounting only for PCR tested positive reporting to the app would have unduly biased the results towards higher severity in the early days. We adopted instead the model developed by Menni et al. (12) that achieved a reasonable performance in prediction of positive cases (ROC-AUC 76%).

## Conclusions

Our data suggest that, in addition to affecting viral transmission, environmental factors also play an important role in already infected patients. Severity of COVID-19 decreased with the onset of spring, which paints a grim picture for the incoming winter and suggest that both disease severity and mortality may increase significantly. Since many hospitals have very dry air in winter, providing humidified air to patients in early stages of the disease may be beneficial. Considering the evident detrimental effect of dry air on our mucosal barrier and its role as the first line of defence against infection (45), in situation of rapidly progressing COVID-19 pandemics it would be essential to actively promote universal humidification of dry air in all public and private heated spaces as well as active nasal hygiene and hydration (46). Humidity should also be monitored in cooled buildings with limited access to outside air, since air-conditioning is also an effective dehumidification and can result in very dry air.

## Data Availability

All data is available upon a reasonable reequest.

## Acknowledgements

We would like to thank all anaesthesiologists and nurses in the Emergency and Intensive Care Unit, as well as all the people who worked at ASST Papa Giovanni XXIII, during the COVID-19 pandemic in Bergamo for their invaluable and brave efforts towards patient’s care. We also want to acknowledge the ROCCO Project (Registry Of Coronavirus Complications) who helped with scientific support, and Dr A. Bonetalli (Epidemiology Office – ASST Papa Giovanni XXIII°, Bergamo, Italy) for her help with data collection. We thank all the medical professionals, nurses and technicians of the ASST GOM Niguarda Hospital who worked hard during the COVID 19 emergency months. Furthermore, we want to acknowledge all the member of the Covid 19 Niguarda Working Group.

We express our sincere thanks to all the participants of the COVID Symptom Study app. We thank the staff of Zoe Global Limited, the Department of Twin Research, and the Clinical & Translational Epidemiology Unit for their tireless work in contributing to the running of the study and data collection.

This manuscript has been released as a pre-print at MedRxiv, https://www.medrxiv.org/content/10.1101/2020.07.11.20147157v2.

---

**Supplementary Figure 1.**
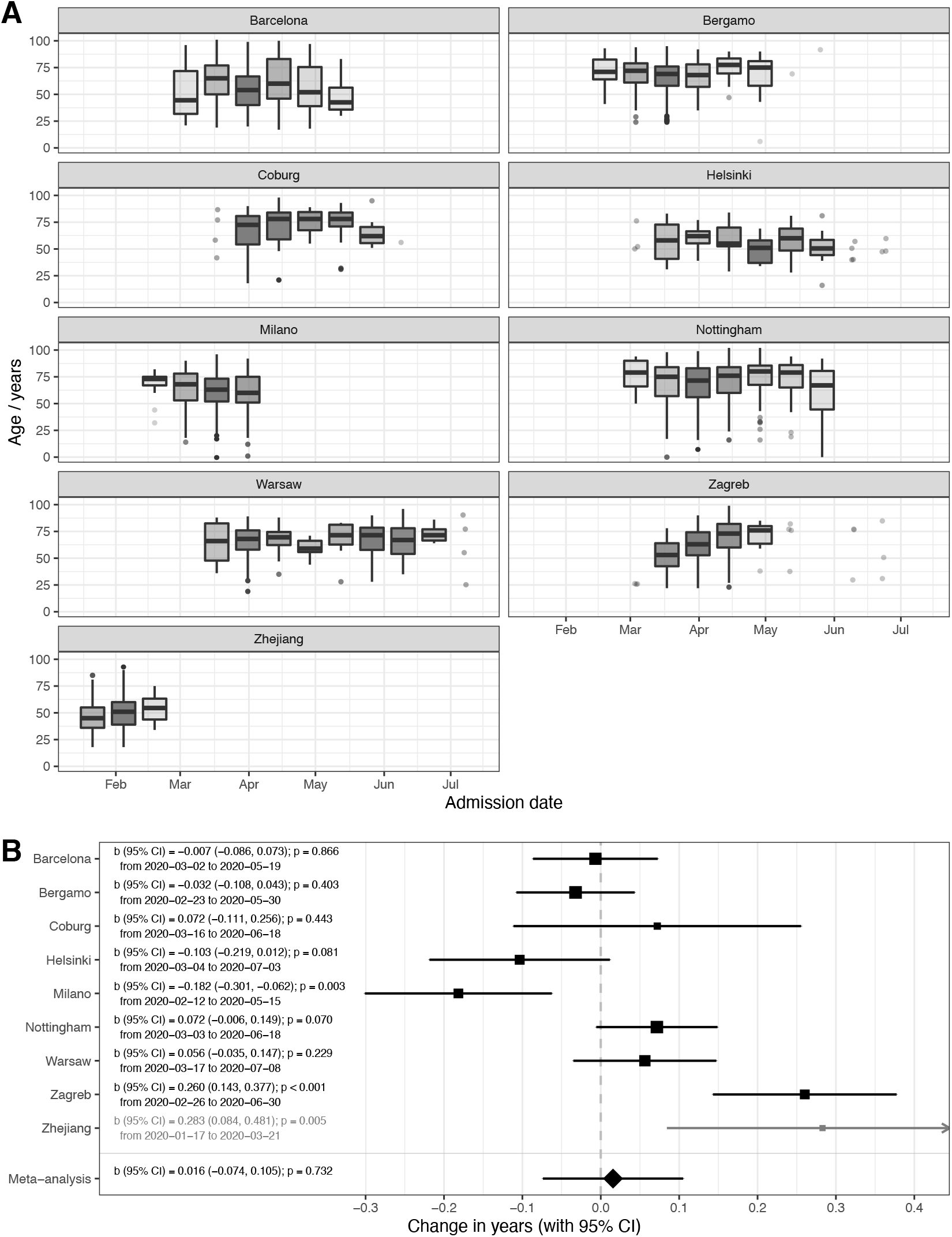
Changes in age of admitted patients with time. A – age boxplots depending on the admission date (grouped in two-week intervals) since the beginning of the pandemics. Lower and upper limits of box present first and third quartile, respectively, and line within the box is median. Whisker lines extend to the minimum and maximum value within ‘inner fence’ defined as 1.5 times interquartile range bellow 1^st^ and above 3^rd^ quartile, respectively. Outliers are presented with dots. If two-week interval had 5 or less values data were presented with dots instead of boxplots; B - Meta-analysis of the effects of admission date on the mortality (presented as odds ratios per one day increase in admission date). In Helsinki there were only 2 deaths and in Zhejiang hospitals 4 deaths, so they were not included in the meta-analysis. OR – odds ratio, CI – confidence interval.

**Supplementary Figure 2.**
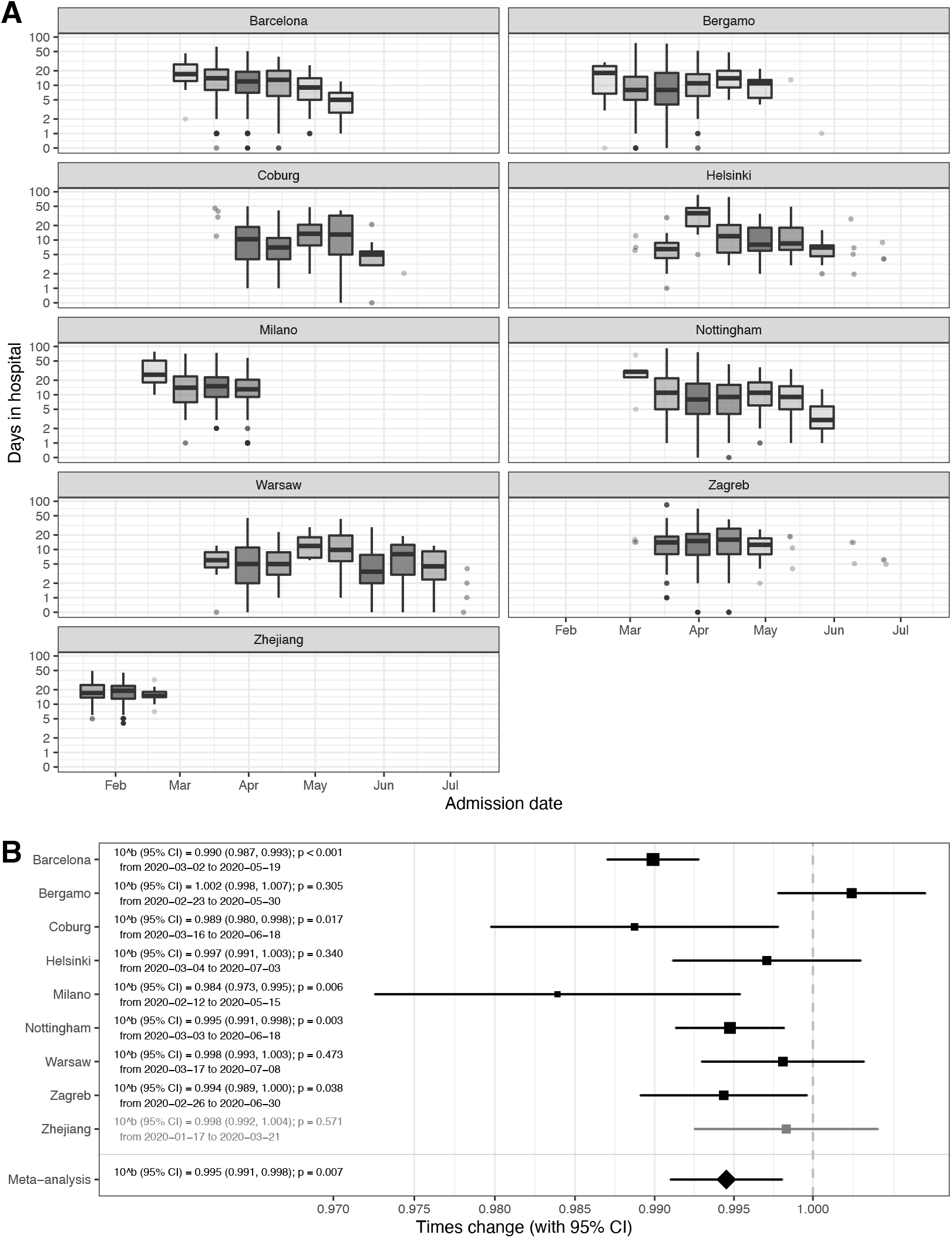
Hospital stay of subjects admitted in hospitals with COVID-19. A - number of days stayed in hospital depending on the admission date (grouped in two-week intervals) since the beginning of the pandemics. Lower and upper limits of box present first and third quartile, respectively, and line within the box is median. Whisker lines extend to the minimum and maximum value within ‘inner fence’ defined as 1.5 times interquartile range bellow 1^st^ and above 3^rd^ quartile, respectively. Outliers are presented with dots. If two-week interval had 5 or less values data were presented with dots instead of boxplots; B - Meta-analysis of the effects of admission date on the hospital stay (presented as times change in duration per each one day increase in admission date). Zhejiang hospital in which all patients were admitted during winter was excluded from the meta-analysis. b – regression coefficient (back transformed), CI – confidence interval.

**Supplementary Figure 3.**
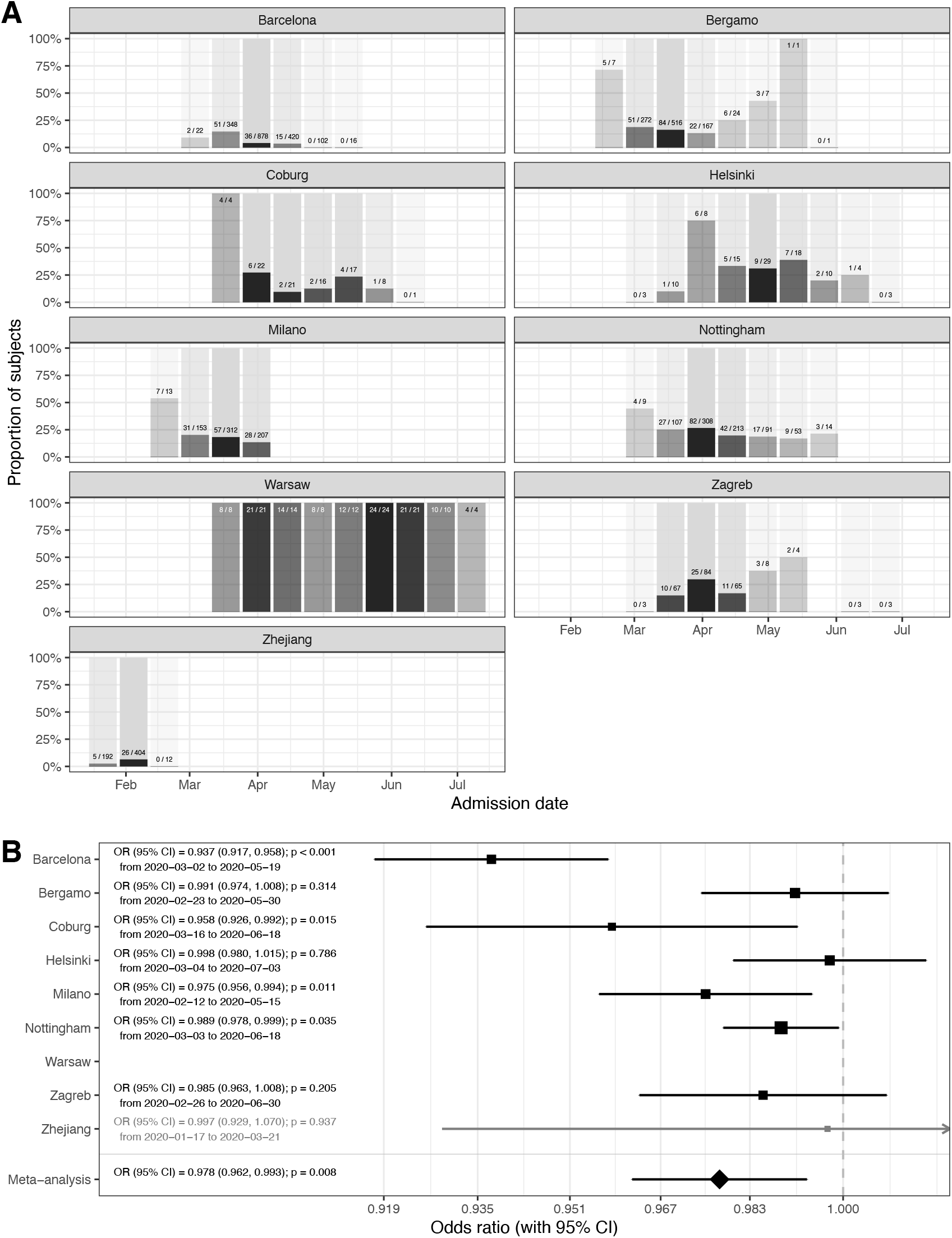
Admission to the Intensive care unit (ICU risk) in people admitted in hospitals with COVID-19. A – proportion of people who were ever admitted to ICU depending on the admission date (grouped in two-week intervals) since the beginning of the pandemics; B - Meta-analysis of the effects of admission date on the ICU admission (presented as odds ratios per one day increase in admission date). Effect of the admission date was not calculated for Warsaw because all subjects were all in ICU. Time period of Zhejiang did not include both cold and warm weather and because of that results were not included in meta-analysis. OR – odds ratio, CI – confidence interval.

**Supplementary Figure 4.**
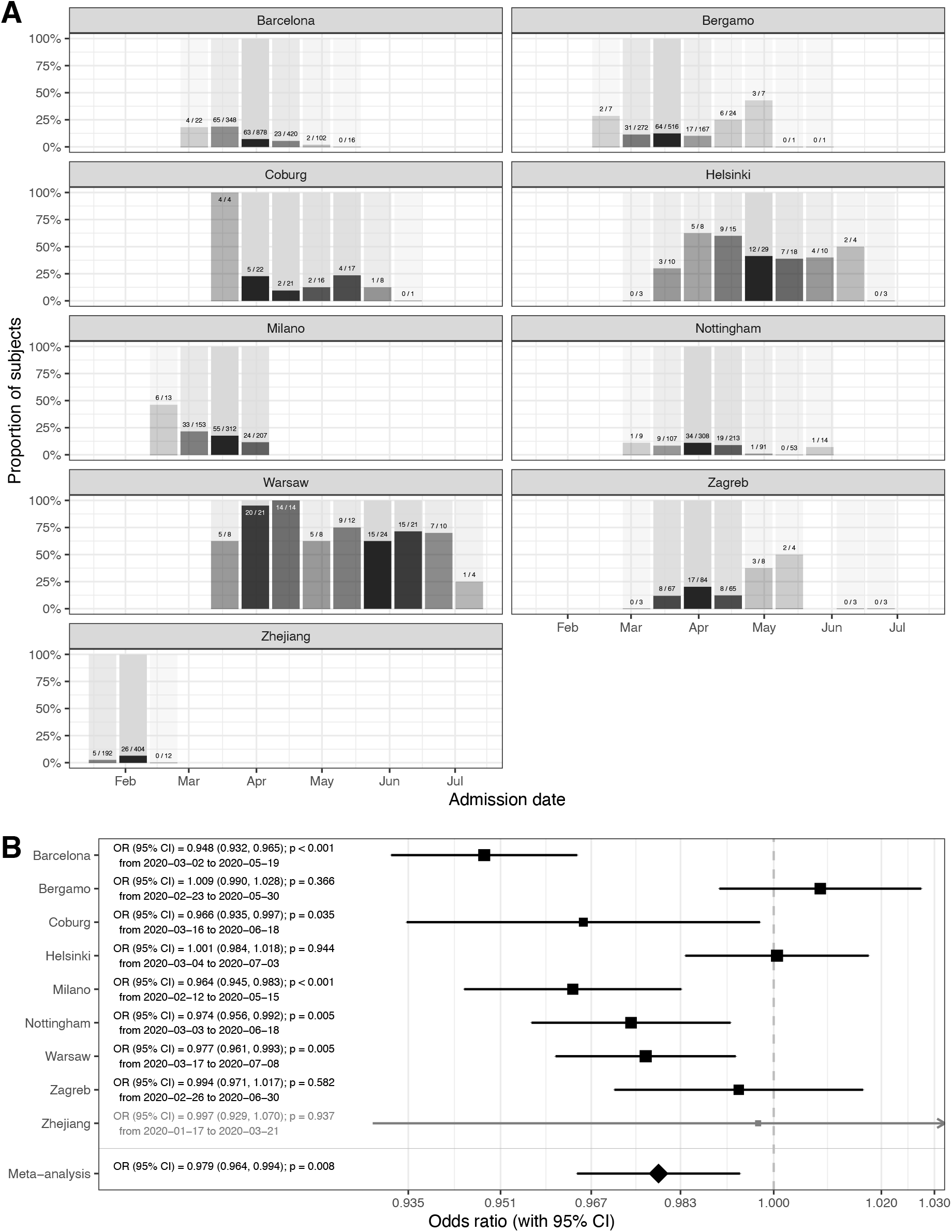
Mechanical ventilation (ventilation risk) in people admitted in hospitals with COVID-19. A – Proportion of people who needed mechanical ventilation depending on the admission date (grouped in two-week intervals) since the beginning of the pandemics; B - Meta-analysis of the effects of admission date on need for mechanical ventilation (presented as odds ratios per one day increase in admission date). Time period of Zhejiang did not include both cold and warm weather and because of that results were not included in meta-analysis. OR – odds ratio, CI – confidence interval.

**Supplementary Figure 5.**
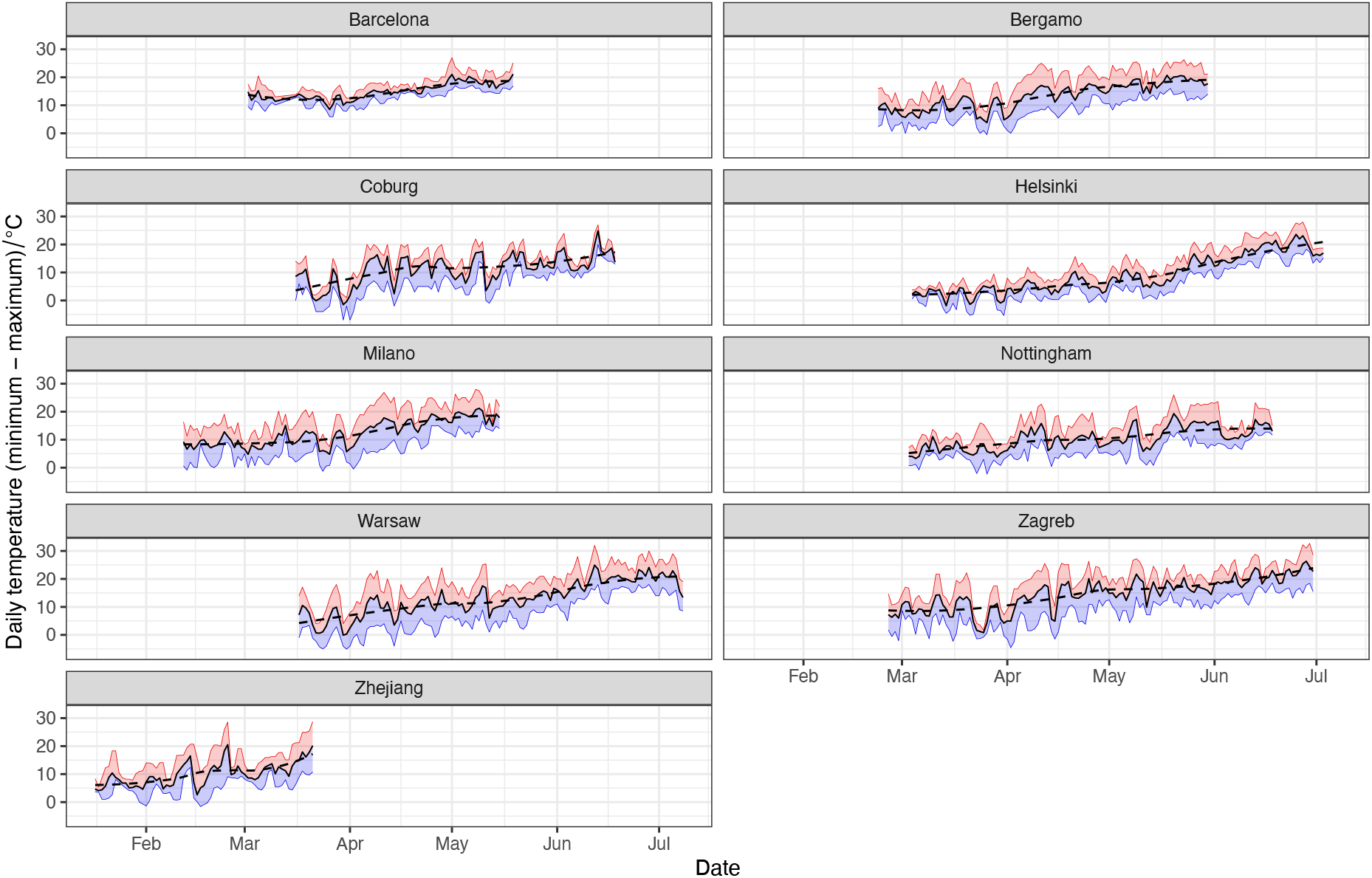
Daily ambient temperatures in the study period. Solid black line – average daily temperature, red line – daily maximum, blue line – daily minimum. Dashed black line – locally estimated scatterplot smoothing of daily average temperature. Data were obtained from the Climate Data Online (National Centers for Environmental Information (NCEI) database): https://www.ncdc.noaa.gov/cdo-web/

